# The New Normal: Delayed Peak SARS-CoV-2 Viral Loads Relative to Symptom Onset and Implications for COVID-19 Testing Programs

**DOI:** 10.1101/2023.05.09.23289735

**Authors:** Jennifer K. Frediani, Richard Parsons, Kaleb B. McLendon, Adrianna L. Westbrook, Wilbur Lam, Greg Martin, Nira R. Pollock

## Abstract

**Background:** Early in the COVID-19 pandemic, peak viral loads coincided with symptom onset. We hypothesized that in a highly immune population, symptom onset might occur earlier in infection, coinciding with lower viral loads.

**Methods:** We assessed SARS-CoV-2 and influenza A viral loads relative to symptom duration in recently-tested adults. Symptomatic participants ≥16y presenting to testing sites in Georgia (4/2022-4/2023; Omicron variant predominant) provided symptom duration. Nasal swab samples were tested by the Xpert Xpress SARS-CoV-2/Flu/RSV assay and Ct values recorded. Nucleoprotein concentrations in SARS-CoV-2 PCR-positive samples were measured by Single Molecule Array. To estimate hypothetical antigen rapid diagnostic test (Ag RDT) sensitivity on each day after symptom onset, percentages of individuals with Ct value ≤30 or ≤25 were calculated.

**Results:** Of 621 SARS-CoV-2 PCR-positive individuals (64.1% women, median 40.9y), 556/621 (89.5%) had a history of vaccination, natural infection, or both. By both Ct value and antigen concentration measurements, median viral loads rose from the day of symptom onset and peaked on the fourth day. Ag RDT sensitivity estimates were 35.7-71.4% on the first day, 63.9-78.7% on the third day, and 78.6-90.6% on the fourth day of symptoms.

In 74 influenza A PCR-positive individuals (55.4% women; median 35.0y), median influenza viral loads peaked on the second day of symptoms.

**Conclusions:** In a highly immune adult population, median SARS-CoV-2 viral loads peaked on the fourth day of symptoms. Influenza A viral loads peaked soon after symptom onset. These findings have implications for ongoing use of Ag RDTs for COVID-19 and influenza.

**Key Points:** In a highly immune adult population, median SARS-CoV-2 viral loads by cycle threshold and antigen measurements peaked on the fourth day of symptoms, with implications for testing practice. In contrast, viral loads for influenza A peaked soon after symptom onset.

## Introduction

Early in the COVID-19 pandemic, SARS-CoV-2 viral loads were observed to peak at the onset of symptoms, and steadily decrease thereafter [1-4]. Subsequently, multiple evaluations of the first commercially-available antigen rapid diagnostic tests (Ag RDTs) indicated that viral loads in symptomatic adults within the first 7 days of symptoms were reliably high enough to be detected in most individuals by the Ag RDTs with strongest performance [5]. Accordingly, initial FDA-approved instructions for use of Ag RDTs utilized only a single test in symptomatic individuals within the first week of symptoms. Early Ag RDT deployment across the United States similarly utilized only one test per symptomatic individual, though the utility of backup molecular testing in those with negative Ag RDT results was simultaneously emphasized [6].

More recently, and particularly during the SARS-CoV-2 Omicron variant surge in early 2022--which overlapped with markedly increased consumer access to home Ag RDTs--increased uncertainty emerged regarding the negative predictive value of Ag RDTs in newly symptomatic individuals and how to guide consumers now in charge of interpretation of their own testing results. Multiple investigations focused on the analytic capability of Ag RDTs to detect the Omicron variant, which appeared to be similar to earlier variants (e.g. [7]). Omicron viral loads were shown to be only slightly lower on average than Delta viral loads [8, 9], and the clinical sensitivity of Ag RDTs (versus polymerase chain reaction (PCR) testing) for Delta vs Omicron variants was also shown to be similar [10]. In parallel, studies of home Ag RDT performance documented improvement of sensitivity with serial testing (e.g. [11]) and in November, 2022, the FDA adjusted recommendations for home Ag RDT use to include guidance to repeat testing in symptomatic individuals 48h after an initial negative test, for a total of at least two tests [12].

Despite the shift over time towards use of serial Ag RDTs in symptomatic individuals to exclude COVID-19, the reasons for the apparent change in clinical performance of Ag RDTs in symptomatic individuals between early and late in the pandemic have not been a major focus of investigation. While there has been much speculation that symptom onset might now be occurring earlier in the course of infection (coinciding with lower viral loads) due to acquired immunity, the current relationship between symptom duration and peak viral load in a highly immune population must be understood in detail to guide Ag RDT testing practice going forward. We therefore sought to assess SARS-CoV-2 viral loads (by PCR cycle threshold (Ct) value and directly-measured antigen concentrations) relative to symptom duration in symptomatic PCR-positive individuals presenting for COVID-19 testing in the past year (after the early 2022 Omicron surge). The use of a multiplexed PCR assay over this timeframe allowed us to also assess the relationship between viral load and symptom duration for influenza A.

## Methods

The Atlanta Center for Microsystems Engineered Point-of-Care Technologies (ACME-POCT) network utilized hospital and community-based COVID-19 testing centers to enroll participants from April 1, 2022 to April 13, 2023 for evaluation of novel viral diagnostic tests under development. Eligible participants were identified consecutively at each study site. Inclusion criteria for the parent study included any person seeking testing for upper respiratory infection. Exclusion criteria for the parent study included patients who were unable to tolerate a nasal swab, were unable to provide informed consent, or had tested positive for SARS-CoV-2 in the two weeks prior to the enrollment date. Following identification, eligible participants for the parent study were approached by study personnel, who obtained informed consent. Clinical electronic case report forms were used to collect demographic and clinical variables, including age, sex, race, COVID-19 related symptoms and time of symptom onset, current health conditions, and exposure, prior infection, and vaccine status. Clinical and demographic variables were collected in a centralized, web-based database (REDCap, Nashville, TN). The study protocol was approved by the Emory Institutional Review Board, Children’s Healthcare of Atlanta and the Grady Research Oversight Committee. For all participants, an FDA-approved RT-PCR SARS-CoV-2 test was administered within 24 hours of study enrollment [anterior nasal swab sample collected with a flocked swab (PurFlock Ultra® Dry Transport System 25-3606-U BT) and diluted in 3.0 mL saline]. For this analysis, only data from patients tested with the Cepheid GeneXpert Xpress SARS-CoV-2/Flu/RSV RT-PCR assay (Cepheid, Sunnyvale, CA, USA) were used. Additional inclusion criteria for our analysis were the presence of symptoms consistent with COVID-19 infection for ≤ seven days at time of testing, age 16 years or older, and positive results of PCR testing for either SARS-CoV-2 or influenza A. Exclusion criteria for this analysis included missing data for the number of days since symptom onset, vaccine status, or prior infection status, and patients with co-detection of SARS-CoV-2 and influenza A or B.

The sample collection date and the self-reported date of symptom onset were used to calculate the number of days of symptoms the patient had experienced at the time of sample collection (with the first day of symptoms designated as Day 0). In a small number of individuals (n=13), a sample collection date had not been recorded, and the date of consent (typically the same day as sample collection, but occasionally the night before sample collection) was used instead.

The Xpert Xpress SARS-CoV-2/Flu/RSV RT-PCR assay (CAT#: XP3COV2/FLU/RSV-10) was performed on fresh anterior nasal swab specimens (see above) within 72 hours of collection per manufacturer’s instructions. The CoV-2 Ct value provided by the Xpert Xpress assay was recorded and used for analysis. For influenza A, Ct values for influenza A target 1 (PA, a phosphoprotein subunit of influenza RNA polymerase); and target 2 (PB2, polymerase basic 2 subunit of influenza RNA polymerase) were recorded and analyzed separately. Samples were frozen (-80C) within 24h after PCR testing.

Nucleoprotein antigen concentrations were measured in the same samples used for PCR testing using the Simoa® SARS CoV-2 N Protein Advantage assay on the Quanterix HD-X platform per manufacturer’s instructions (Quanterix; Billerica, MA). Samples had been frozen once prior to antigen measurement and were tested within 12h of thaw. Samples which generated values that were above the highest calibrator were diluted to generate a final measurement. For this analysis, any antigen concentration reading that was below either the lower limit of quantification or the lower limit of detection on the instrument was recoded to 0 pg/mL, and any sample with an antigen concentration reading above quantifiable range that had not undergone dilution testing to obtain an actual concentration was excluded from this analysis. Any antigen concentration value that was missing was excluded from this analysis. For normality and to account for naturally occurring 0’s, antigen concentration (pg/mL) was natural log transformed(n+1).

Days since symptom onset and Ct values or log transformed antigen concentrations were plotted using box-and-whisker plots to represent median/IQR. Counts of each symptom present were plotted using stacked bar charts. In order to estimate the hypothetical performance of Ag RDTs in this population, we also calculated the percentage of participants with a Ct value less than or equal to 30 or less than or equal to 25. All plots and tables were generated using the ggplot2, dplyr, and r2rtf packages in R Statistical Software (v4.2.0; R Core Team 2023; Vienna, Austria) [13-15].

## Results

There were 621 participants that met our inclusion criteria; 64% were women and the median age was 41 years. Of the 621 individuals, 556 (89.5%) had a history of SARS-CoV-2 vaccination, natural infection, or both, confirming a high level of SARS-CoV-2 immunity in the study population. Supplementary Figure 1 plots the symptom distribution in these participants; the symptoms with the highest prevalence were cough (83.3%), rhinorrhea/runny nose/congestion (77.3%), and sore throat (71.8%). Figure 1A shows the SARS-CoV-2 Ct values from testing of nasal swab samples from these symptomatic PCR-positive participants plotted by the duration of symptoms at the time the sample for PCR was collected (day 0 = the first day of symptoms). Median Ct values hit their nadir (consistent with peak viral loads) on the fourth day of symptoms and then trended up on the following days. Plotting of paired antigen concentrations (measured in the same samples that received PCR testing; Methods) confirmed the same pattern, with median antigen concentrations rising from the day of symptom onset and peaking on the fourth day of symptoms (Fig 1B).

**Figure 1.**
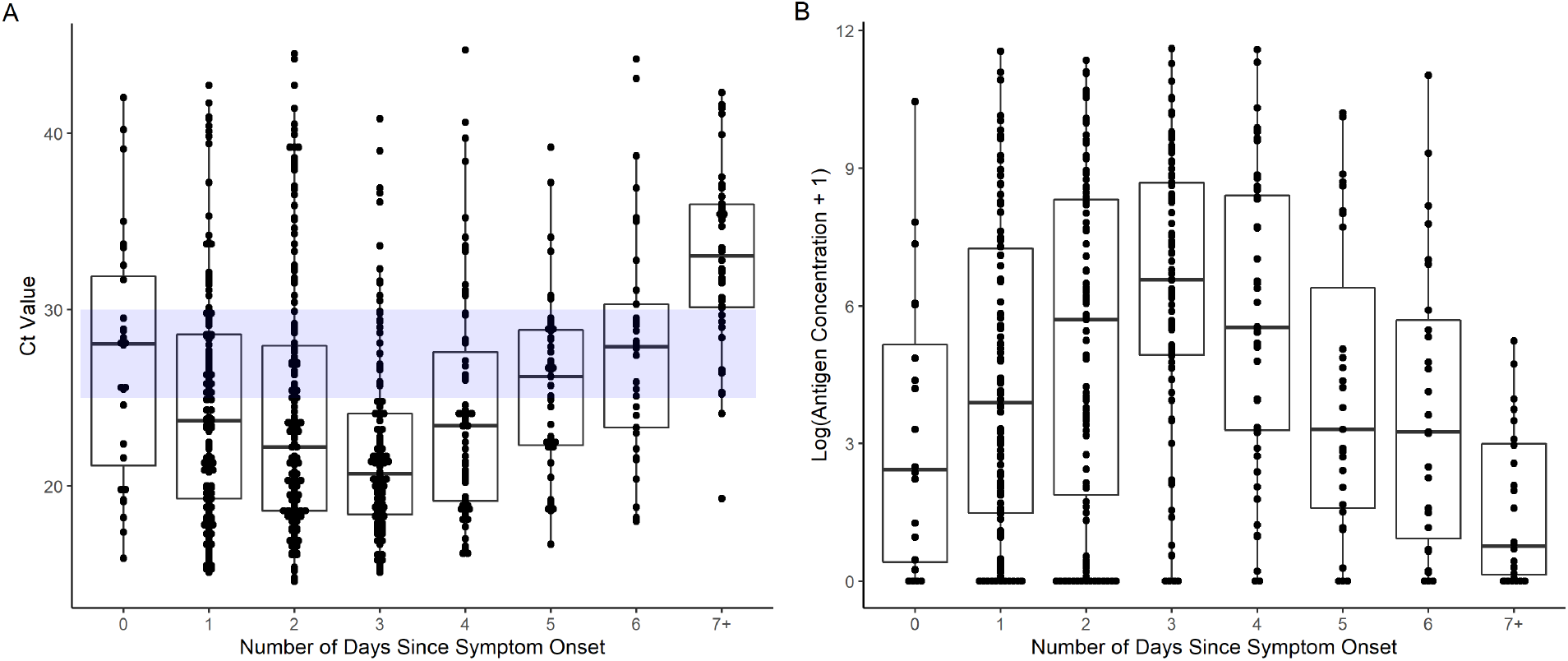
SARS-CoV-2 Ct values (A) and paired log-transformed antigen concentrations (B) measured in anterior nasal swab samples plotted by days since symptom onset for PCR-positive symptomatic adults (day 0 = the first day of symptoms). The blue bar in Figure 1A outlines the window between Ct of 30 and Ct of 25. Ct, Cycle threshold.

To estimate the % of PCR-positive individuals who might be expected to test positive by Ag RDT on each day after symptom onset, the % of participants with a Ct value ≤ 30 (a threshold chosen to represent the most sensitive Ag RDTs commercially available, based on internal data [16]) versus ≤ 25 (representing the least sensitive Ag RDTs available) on each day were calculated (Table 1; see also blue shading, Figure 1A). On the first day of symptoms, sensitivity of rapid testing (compared to PCR) was estimated to range from 35.7-71.4%, and on the fourth day of symptoms, from 78.6-90.6% (Table 1). Estimated sensitivity on the third day of symptoms (notably, corresponding to the timing of the FDA recommendation to repeat testing 48h after an initial negative test, [12]) was 63.9-78.7%. For all participants within the first 7 days of symptoms, sensitivities ranged from 60.8-82.2%.

**Table 1.** SARS-CoV-2 Cycle Threshold (Ct) values, number/percent of samples with Ct values no more than Ct 25 or Ct 30, and nucleoprotein antigen concentrations, grouped by days since symptom onset.

| Days Since Symptom Onset | Ct, median (IQR) | Ct $\leq 25$, n (%) | Ct $\leq 30$, n (%) | Ag conc (pg/mL), median (IQR), n |
| --- | --- | --- | --- | --- |
| 0, n=28 | 28.0 (21.2-31.9) | 10 (35.7) | 20 (71.4) | 10.4 (0.5-199.4), n=20 |
| 1, n=136 | 23.7 (19.3-28.6) | 76 (55.9) | 113 (83.1) | 48.1 (3.4-1407.9), n=106 |
| 2, n=155 | 22.2 (18.6-28.0) | 99 (63.9) | 122 (78.7) | 298.1 (5.6-4121.6), n=111 |
| 3, n=117 | 20.7 (18.4-24.1) | 92 (78.6) | 106 (90.6) | 714.2 (138.1-5939.2), n=87 |
| 4, n=67 | 23.4 (19.1-27.6) | 45 (67.2) | 54 (80.6) | 251.8 (25.9-4509.0), n=48 |
| 5, n=43 | 26.2 (22.3-28.9) | 19 (44.2) | 37 (86.0) | 26.5 (3.9-1205.5), n=31 |
| 6, n=33 | 27.9 (23.3-30.3) | 11 (33.3) | 24 (72.7) | 24.8 (1.6-302.0), n=27 |
| 0-6, n=579 | 23.1 (19.1-28.4) | 352 (60.8) | 476 (82.2) | 156.7 (6.7-2890.1), n=430 |
| 7+, n=42 | 33.0 (30.1-36.0) | 2 (4.8) | 10 (23.8) | 1.1 (0.2-19.1), n=28 |

Ct, cycle threshold; IQR, interquartile range; Ag, antigen; Conc, concentration

There were 74 symptomatic adults who presented for testing and were PCR-positive for influenza A. These participants had a median age of 35 years and 55% were women. No influenza vaccination data were available. Ct value distributions (for the two influenza A targets from the Xpress assay) plotted by duration of symptoms at the time of testing are shown in Figure 2. Median/IQR trends (Fig 2, Supplementary Table 1) for both influenza A targets suggest that in this population, influenza A viral loads peaked shortly after symptom onset--unlike the trend observed for SARS-CoV-2. Of the 21 participants positive for influenza A on the second day of symptoms (day 1), 85.7% and 76.2% had Ct values </= 30 and </= 25, respectively (Supplementary Table 1).

**Figure 2.**
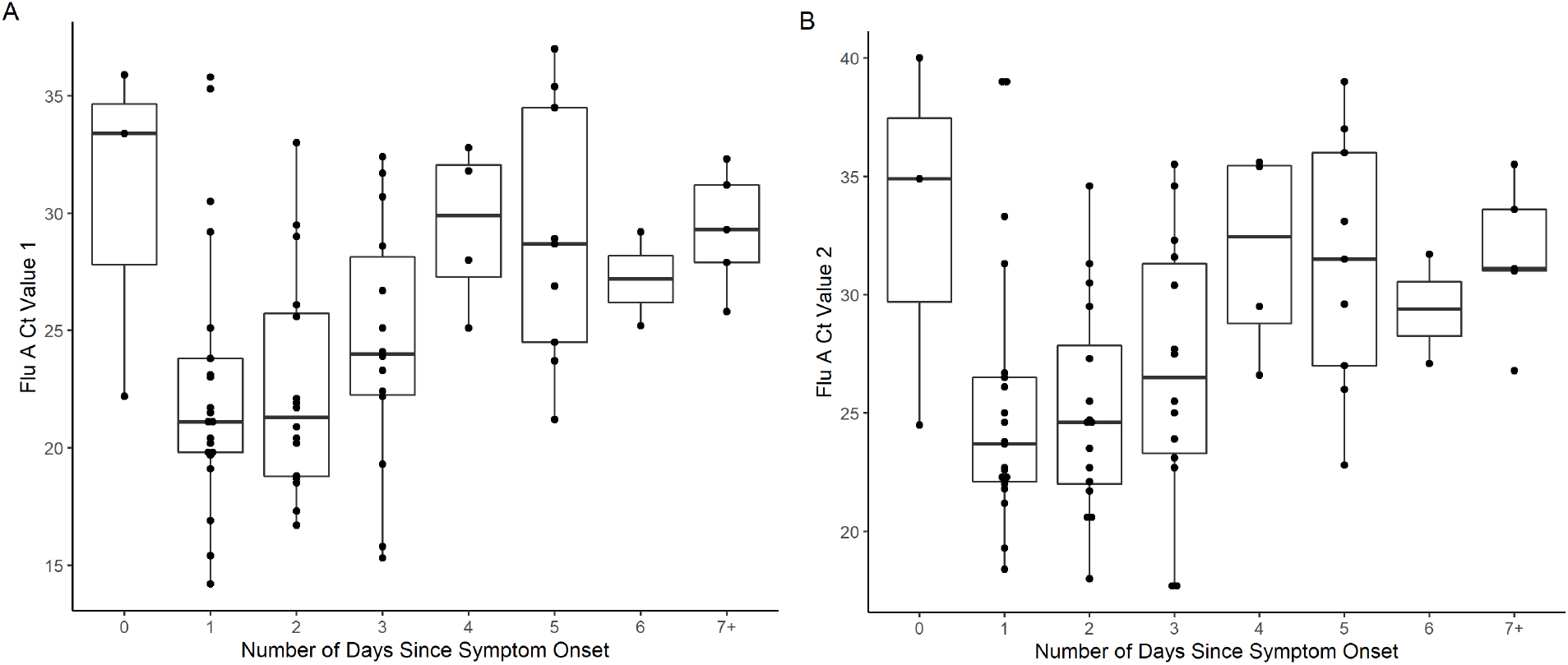
Influenza A Ct values measured in anterior nasal swab samples plotted by days since symptom onset for PCR-positive symptomatic adults (day 0 = the first day of symptoms). A, Ct value for influenza A target 1 (PA, a phosphoprotein subunit of influenza RNA polymerase); B, Ct value for influenza A target 2 (PB2, polymerase basic 2 subunit of influenza RNA polymerase. Ct, cycle threshold.

## Discussion

Our data suggest that the relationship between SARS-CoV-2 Ct value distributions (as a well-established proxy for viral load distributions) and the timing of symptom onset in a highly immune population is very different than the relationship between these parameters observed early in the pandemic—a finding with major implications for testing practice going forward.

Early in the pandemic, multiple studies showed that viral loads were highest at the time of symptom onset and then declined steadily thereafter [1-4]. The sensitivity of rapid antigen tests within the first 7 days of symptoms was also quite high in multiple studies performed early in the pandemic, with overall sensitivities (versus PCR) >90-95% in the first week of symptoms in multiple studies (e.g. [17-20]). In short, early in the pandemic, a single negative antigen test had reasonable negative predictive value early in the course of symptoms.

Now, in this highly immune population presenting after the early 2022 omicron surge, our data suggest that viral loads appear to peak four days after the onset of symptoms. Predicted rapid antigen test sensitivity on day 4 peaked at 78.6-90.6%, while overall predicted sensitivity within the first 7 days of symptoms was only 60.8-82.2%. Our choice of Ct cutoffs of 25 and 30 for predictions of hypothetical rapid antigen test sensitivity were based both on the numerous studies assessing antigen test sensitivity (versus a variety of PCR assays) at these cutoffs (e.g. [17-20]) and on internal data evaluating the sensitivity of commercial rapid antigen tests versus the Cepheid Xpert Xpress assay at point-of-care and using omicron swab samples in transport media [16]. In this study, trends observed for directly-measured antigen concentration distributions very closely mirrored Ct value trends, providing additional support for the antigen test sensitivity predictions and suggesting that the close correlation between Ct value and Ag concentration observed in samples from early in the pandemic remains [21].

This delayed peak relative to the onset of symptoms has been observed in other studies conducted later in the pandemic [22-24], but the implications of this finding for testing practice—in particular, home antigen testing practice—have not been sufficiently highlighted. Hay et al [22] evaluated and modeled PCR test results in a cohort of mostly young healthy men (18% unvaccinated) and observed a median 2-3 day interval between onset of symptoms and peak viral loads (nadir Ct values) for both delta and omicron infections, but did not present Ct value distributions by day of symptoms to inform Ag RDT use. Kandel et al [23] evaluated serial testing data for 37 vaccinated adults with incident omicron infection and found that viral load peaked 3 days after symptom onset. Interestingly, this shift to a later peak viral load relative to symptom onset may have been observed even by the second year of the pandemic, given that a study done in the first half of 2021 [24] (pre-delta variant, with 86% of participants unvaccinated) using serial home Ag RDT testing already demonstrated that Ag RDT sensitivity in adults and children with PCR-confirmed infection peaked 4 days after symptom onset. We note that none of these prior studies included antigen concentration measurements, and our finding that the curves for Ct value and Ag concentration data were so similar suggests that the kinetics of RNA and protein clearance from infected tissue may be similar.

While the observation of a delayed peak (relative to onset of symptoms) compared to Ct value distributions observed early in the pandemic is consistent overall with current FDA recommendations to repeat rapid antigen testing 48h after an initial negative test [12], our data in combination with others’ suggest that symptomatic individuals testing positive for SARS-CoV-2 by PCR currently may not reliably test positive on a rapid antigen test until the 3^rd^ or 4^th^ day of symptoms. Though testing negative on an antigen test suggests that the person is less likely to be viral culture-positive (and presumably infectious) at that moment than someone who tests positive [25], we note that our data indicate that viral load would be expected to be rising, and not falling, in the window after that early false negative test. Additionally, we note that samples containing the omicron variant have been observed to be culture-positive at lower viral loads than samples containing the delta variant [26], suggesting that the reassurance provided by negative antigen tests may be even less now than earlier in the pandemic. Individuals who test negative on antigen tests on the first or second day of symptoms—or even on the third-- and who remain symptomatic need to clearly understand that COVID has not been excluded and that they should take precautions around others through the fourth day of symptoms, including masking when feasible. Ideally, another test would be done on the fourth day of symptoms. Our findings also have implication for test-to-treat programs that combine home Ag RDT use with prescriptions of antiviral treatment [27], given that treatment is optimally given as early in the course of infection as possible.

Our additional finding that influenza A viral loads were highest early in the symptom course— similar to the trends observed early in the COVID pandemic, and very different than the current trends we observed for SARS-CoV-2 infection--has important implications for test development and testing practice, though we note that we were unable to assess the impact of prior vaccine- or infection-based immunity on these findings. Multiple manufacturers are currently engaged in the development of multiplexed tests capable of simultaneous detection and distinction of SARS-CoV-2 and influenza viruses, and discordance in the timing of peak viral loads for the different viruses means that multiplexed tests will have major variance in negative predictive values at different time points within the course of one illness. Consumers, particularly those performing multiplexed tests at home, will need to be made aware of this variation so that the meaning of negative results can be more clearly understood.

Limitations of our study include that we utilized PCR Ct values as a proxy for viral loads. However, given that our goal was to assess trends in viral load kinetics relative to days of symptoms, utilized one PCR assay for the entire study, and were able to directly assess antigen concentrations in parallel with Ct values, we do not believe that this is an important limitation. We acknowledge that our population-level analysis may not perfectly represent viral load kinetics at the per-patient level, though our findings are consistent with serial testing data [23]. We did not have enough participants to clearly evaluate the impact of varying forms of immunity (vaccine, prior infection, or both) on Ct value trends, but given that almost everyone presenting for testing in the study window had some known form of immunity, we think that our results are generalizable to the larger US population at this point in the pandemic. Similarly, we did not confirm the variants responsible for the infections in our study population, though local/national circulation of Omicron variant strains was well-established during the study timeframe. Our influenza data are limited by the relatively small sample size and the lack of information on vaccination, and we recognize that we are not able to clearly predict the implications for influenza A Ag RDT testing of the Ct value trends we observed.

In summary, our data remind us that viral load kinetics relative to duration of symptoms can change over time with exposure to an initially novel virus, and that data collected early in a pandemic should not be assumed to still apply in the months to years that follow. Capture of duration of symptoms data at the time of testing is both simple and high yield, and should be incorporated into testing programs so that these trends can be monitored and continue to guide testing practice going forward.

## Data Availability

All data produced in the present work are contained in the manuscript

## Author Contributions

J.K.F., R.P., A.L.W., and N.R.P. conceived and designed the analysis. K.B.M. contributed to data generation. J.K.F., R.P., K.B.M., A.L.W., W.L, and N.R.P. analyzed and/or interpreted the data. N.R.P., R.P., and J.K.F. drafted the manuscript. Critical revisions to the manuscript were made by all members of the study group. All authors had full access to all the data in the study and had final responsibility for the decision to submit for publication.

---

**Supplementary Figure 1.**
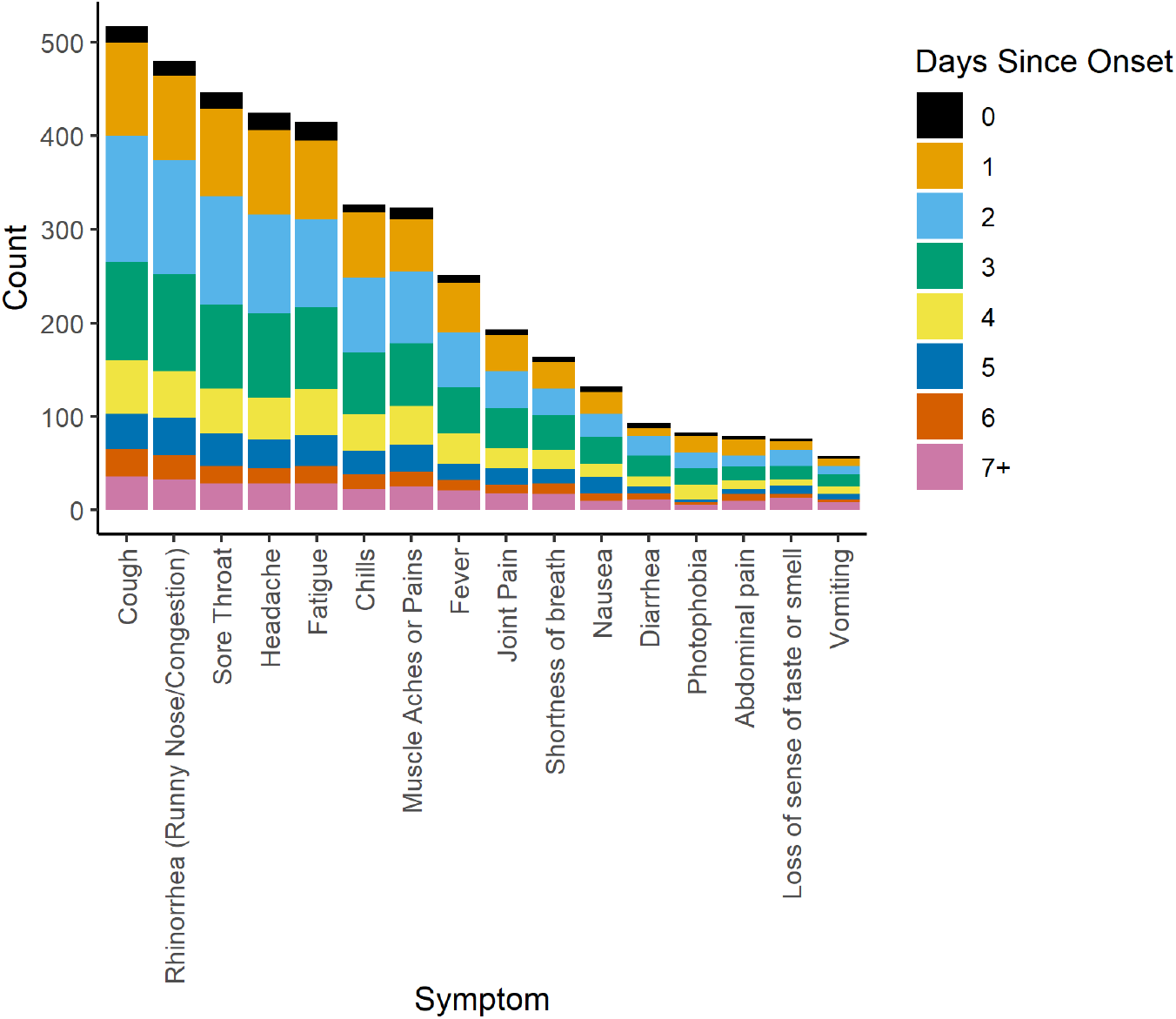
Presence of each of 16 recorded symptoms in the study population, grouped by days since symptom onset.

**Supplementary Table 1.**
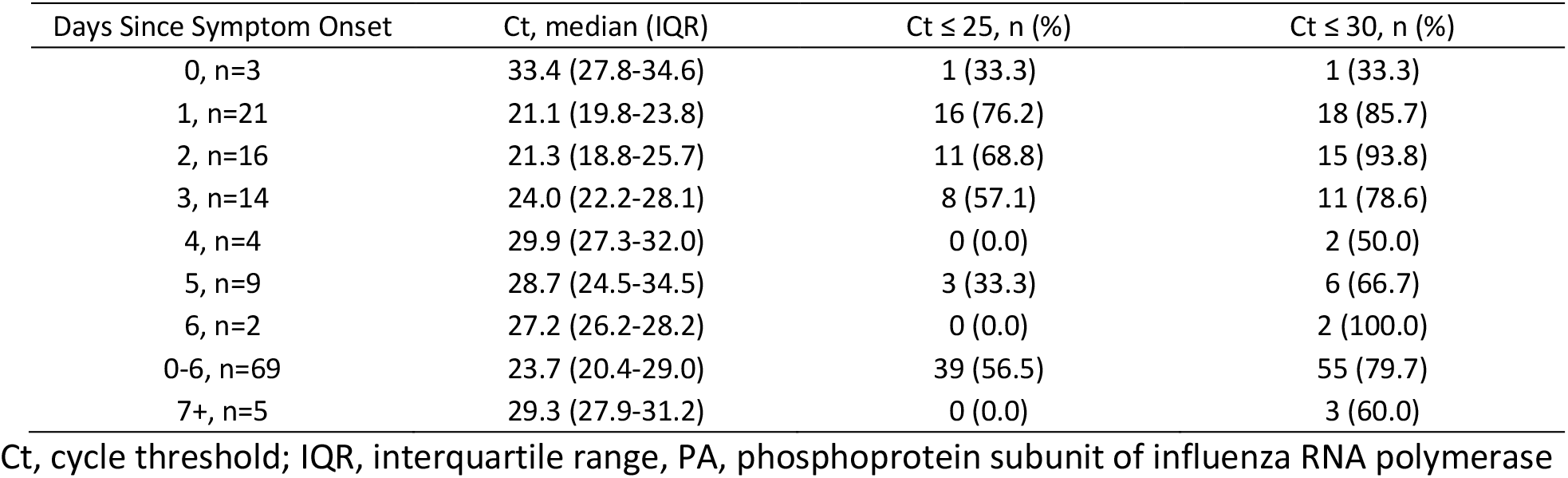
Influenza A Cycle Threshold (Ct) values (PA Target) and number/percent of samples with Ct values no more than Ct 25 or Ct 30, grouped by days since symptom onset.

